# The Acceptability of Three Co-Created Peer Support Interventions for People Living with Leprosy Reactions in Indonesia: A Mixed-Methods Pilot Study

**DOI:** 10.64898/2026.06.10.26355364

**Authors:** Annisa Ika Putri, Stephen L. Walker, Regitta Indira Agusni, Medhi Denisa Alinda, Bagus H. Kusumaputra, M. Yulianto Listiawan, Marjolein B.M. Zweekhorst, Ruth M.H. Peters

**Affiliations:** Athena Institute, Vrije Universiteit Amsterdam, Netherlands; Department of Dermatology Venereology & Aesthetic, Dr. Soetomo General Academic Hospital, Surabaya, Indonesia; Department of Clinical Research, Faculty of Infectious and Tropical Diseases, London School of Hygiene and Tropical Medicine, London, United Kingdom; Department of Dermatology Venereology & Aesthetic, Faculty of Medicine, Universitas Airlangga, Indonesia; Department of Dermatology Venereology and Aesthetics, Universitas Airlangga Hospital, Indonesia; Leprosy Study Group, Institute of Tropical Disease, Indonesia

**Keywords:** Peer counselling, Telesupport groups, Participatory video, Leprosy reactions, Peer support, Co-creation

## Abstract

**Background:** Leprosy reactions (LR) are immune-mediated complications associated with disability, emotional distress, and social isolation. We identified a gap in affected-individual-informed interventions that aim to improve the management of LR in healthcare settings. To address this gap, we assessed the acceptability of three peer-support interventions co-created with people affected by LR in Indonesia.

**Methods:** Using an interactive learning and action approach, we co-created peer counselling, telesupport groups, and participatory video interventions which were piloted in an urban hospital and 13 rural community clinics. A mixed-methods design was applied with interviews, focus group discussions, and pre–post assessments involving four participant groups. Data were analyzed thematically using an acceptability framework.

**Results:** One hundred participants were enrolled, and 92 completed the pilot intervention between November 2022 and July 2023. Qualitative findings showed that all interventions were acceptable. Peer counselling provided emotional reassurance through shared experiences and was perceived as trustworthy and supportive. Perceived burdens differed by setting, with time constraints in urban facilities and geographical barriers in rural clinics. Knowledge improved significantly among participants of peer counselling and telesupport groups in rural settings. Telesupport groups facilitated connection, information exchange, and continuity of care. Digital access and literacy limited participation for some, particularly in rural areas. The participatory video was perceived as reassuring and informative. Improvements in knowledge, attitude, practices, and mental well-being domain scores were observed among urban participants, but responses in rural settings showed less change. Participants and co-implementers reported increased self-efficacy, participants’ confidence to perform required behaviors within peer support interventions, with effects shaped by intervention and setting.

**Conclusions:** The three co-created peer-support interventions were acceptable for individuals with LR in diverse healthcare settings. These outcomes highlight the importance and effectiveness of selective, and context-sensitive implementation of one or more peer-support modalities.

**Author summary:** People affected by LR may experience severe physical, psychological, and socioeconomic impacts, including pain, nerve damage, emotional distress, stigma, and loss of income and social roles[1,2]. LR place a significant burden on leprosy services as they require prolonged treatment and may adversely affect the lives of those affected. Our findings from an earlier study revealed a need for holistic interventions to improve the management of LR in Indonesia[1,2]. Most studies have focused on treatment of LR, with less exploration of tailored psychosocial approaches in healthcare settings for affected individuals. Peer counselling has been effective in reducing stigma and improving quality of life in various health contexts, including HIV, mental health, and leprosy in general. Telesupport groups are effective for remote health support. Participatory video has shown promise in empowering marginalized groups and reducing stigma. However, the three interventions have not yet been developed for individuals affected by LR.

To our knowledge, this is the first study to develop and pilot tailored peer counselling, telesupport, and participatory video for individuals experiencing LR in hospital or community clinic settings. We demonstrated that peer support interventions were acceptable in all the healthcare facility settings. Interventions were reported by participants to improve knowledge, self-motivation, and treatment adherence through sharing experiences. There were measurable improvements in knowledge, attitude, and willingness to disclose their leprosy diagnosis in particular for urban participants. Observed variation in facilitating factors of intervention’s acceptability across study sites highlights the influence of infrastructure, digital access, literacy, and healthcare capacity on intervention uptake and impact.

This study provides evidence that peer-support approaches complement clinical management of LR and improve stigma-sensitive care. Integrating these interventions into routine leprosy services could strengthen patient education, motivation, and social reintegration. Implementation should include peer counsellor training, investment in digital infrastructure, and sustained collaboration between health workers and affected stakeholders to promote holistic, person-centered care in leprosy and other neglected tropical diseases.

## Introduction

Leprosy is a significant public health problem in Indonesia. Indonesia remains one of the 23 World Health Organization (WHO) global priority countries and consistently reports the third highest number of new cases annually[3]. Indonesia is a major contributor to the regional burden accounting for approximately 82% of newly detected cases in Western Pacific with 14,698 new cases reported to WHO in 2024[3]. In Indonesia, over 90% of these cases were multibacillary (MB) of whom 869 (5.9%) had disability at diagnosis and 1,420 (9.7%) were children[4].

The burden of leprosy extends beyond infection. Approximately 60% of individuals with leprosy experience severe immune-mediated inflammatory episodes called leprosy reactions (LR)[5]. LR may occur before, during, or after completion of multi-drug therapy (MDT) and cause substantial physical and psychological impact[1]. LR are classified as Type-1 reactions (T1R) and Type-2 reactions or erythema nodosum leprosum (ENL), and can lead to irreversible nerve damage, pain, disability and may necessitate hospitalization[5,6].

T1R are characterized by acute inflammation in pre-existing skin lesions and/or nerves, whereas ENL presents with recurrent crops of painful, erythematous nodules often accompanied by fever and systemic inflammation[7]. LR often recur and require long-term management. They may complicate adherence to MDT, and undermine confidence in healthcare workers and care organizations[8,9]. LR data reported to WHO is incomplete and only reflects LRs present at the time of leprosy diagnosis (and reporting). T1R reactions occur in approximately 20%-40% of leprosy-affected individuals and ENL occurs in 50% of individuals with lepromatous leprosy[10–13].

LR and its treatment may be associated with stigma, discrimination, and social exclusion because of visible skin changes, disrupted social roles, and self-isolation[1,14]. For example, common cutaneous manifestations of LR on skin (inflammatory nodules, pustules, and ulcers) are often more noticeable than other leprosy symptoms without reactions and may lead to stigma[1,15]. High-dose clofazimine often used to treat ENL, causes skin discoloration leading to self-isolation and discrimination[1,16]. Painful reactions restrict mobility, making it challenging to work or manage household tasks[1]. Family members may blame their affected relative for their limitations reinforcing moral judgement and rejection[1,17]. Furthermore, prolonged hospitalization interrupts school attendance, employment and household roles. This leads to educational delay, unemployment, reduced income, and perceived failure as a breadwinner or caregiver, which heightens both anticipated and experienced stigma[1,15,18].

LR contribute to emotional distress, anxiety, and depression[1]. However, psychosocial care for individuals with LR is underdeveloped and poorly integrated into routine leprosy services in both hospital and community-based settings[2]. Resource-limited settings lack trained personnel and psychosocial programs to support the management of LR[2]. We identified potential benefits from social and emotional support (in addition to clinical care) such as community-based, person-centered approaches to cope with the long term physical and psychosocial aspects of LR[1,2]. Indonesian participants in our previous study suggested peer-support would help in dealing with the impact of LR by enabling the sharing of common experiences and providing mutual comfort and support[1].

Several interventions have shown promise or demonstrated effectiveness in improving the care and psychosocial well-being of individuals affected by leprosy and other health conditions. Trained peer counsellors with lived experience provide emotional and practical support, reducing stigma and improving treatment adherence across stigmatized conditions such as leprosy, HIV, and mental health disorders[19,20]. Peer counsellors foster empathy and mutual understanding[21,22]. Telecommunication technologies have emerged as valuable platforms for providing remote, group-based psychosocial support particularly since the COVID-19 pandemic[23,24]. Telesupport groups are deemed appropriate to facilitate real-time discussions on self-care, treatment experiences and coping strategies among individuals affected by chronic conditions such as dementia, diabetes mellitus, cancer and multiple sclerosis[25–34]. Telesupport groups reduce feelings of isolation and geographical and social barriers to care[28,31,32]. Participatory video (PV) offers a collaborative means for individuals to share their personal experiences of recovery, empowerment, communication, and stigma reduction through the video-making process[35,36]. PV has been applied widely with stigmatized groups across health research, including leprosy-affected individuals[37–41].

Peer counselling and PV have demonstrated effectiveness among people affected by leprosy but evidence on the acceptability of these interventions among individuals with LR is limited. Telesupport groups in leprosy have not been explored despite its increasing use in other chronic and stigmatized conditions. Understanding whether these approaches are acceptable in real-world hospital and community clinic settings is particularly important in a high burden leprosy endemic country such as Indonesia. We aimed to assess the acceptability of peer counselling, telesupport groups, and PV among individuals affected by LR attending hospital and community clinic settings in Indonesia.

## Methods

### Study design

Our participatory study adopted the interactive learning and action approach with a mixed-methods design. The research team consisted of three research assistants and a junior researcher supervised by five senior researchers. People affected by LR were the main stakeholders and co-implementers. Eight co-implementers from urban and rural setting (four from each) acted as peer counsellors, facilitators of the telesupport group, and participatory video makers. A larger group of 12 people affected by LR and the eight co-implementers, together with the research team and four healthcare professionals, were involved in an iterative process to collaboratively design the proposed interventions, plan the pilot implementation, and make any necessary adjustments to the piloted interventions. Adjustments were made to consistently incorporate the needs of people with LR, considering the time and resources available.

### Study sites and participants

Two study sites were purposively selected in East Java, one of Indonesia’s leprosy-endemic regions. The study took place in Dr. Soetomo General hospital in Surabaya, an urban referral setting, and 13 community clinics that provide leprosy and other primary care services such as TB, HIV, maternal and childcare in Bangkalan, a rural district. The eight co-implementers were recruited from 33 individuals who participated in a previous study exploring improvement in the management of LR[1]. Eligibility criteria for co-implementers included completion of LR treatment and availability to carry out the role. All co-implementers received training on the three interventions, regardless of which intervention they facilitated, as outlined in the summary of the training module (Supplementary File 1). Intervention participants were recruited purposively by the research team from November 2022 to July 2023. The eligible participants were adults aged 18 years or older and users of LR care services who were recruited during outpatient visits. Accompanying family members (FM) were recruited after being proposed as potential participants by individuals with LR.

### Pilot interventions

#### Peer counselling

Peer counselling consisted of five one-to-one sessions timed to coincide with ‘LR-affected participants’ routine outpatient visits at the clinic. The individual session was scheduled based on availability and agreement of the co-implementer and participant. Each session lasted 30–45 minutes and was delivered by the trained co-implementers under the supervision of the study team. Session content covered knowledge of leprosy and LR, emotional coping, family support, and creating a personal action plan to reduce stigma.

#### Telesupport groups

Telesupport groups were conducted via text-based discussions in an “urban” and “rural” WhatsApp group. Each group comprised individuals affected by LR and was facilitated by four co-implementers who were assigned based on their place of residence (urban or rural) to align facilitators’ and participants’ local context. The groups were monitored for eight weeks. Discussions were semi-structured with one topic introduced each week including information on leprosy and LR (symptoms, risk factors, triggers, and treatment), personal experiences of treatment adherence, and challenges in accessing healthcare services. Co-implementers initiated conversations by sharing lived experiences and moderated peer exchange, while clinicians were consulted when medical clarification was required. Most discussions were conducted through text messages. In addition, audio messages and digital photographs were allowed for group members unable to read. Each participant had equal opportunity to give their opinion within the group. The same co-implementers who delivered peer counselling also facilitated the telesupport groups, adapting their role from one-to-one support to digital group facilitation. Further details on discussion topics and facilitation are provided in Supplementary file 2.

#### Participatory video

One participatory video was co-created with the eight co-implementers (who were the same individuals involved in peer counselling and telesupport groups). The 9-minute video was semi-scripted and featured testimonial-style storytelling in an interview format. In the video, co-implementers shared their lived experiences of LR, including their treatment journeys and coping strategies. It was produced in Javanese and Madurese and subtitled in Bahasa Indonesia. The video was viewed individually on a research assistant’s smartphone or laptop by LR-affected individuals and their FM in the same waiting area. Individual viewing was used because group video screening was not feasible due to space and scheduling constraints. Each participant viewed the video once together with either a research assistant or a co-implementer.

The pilot interventions were monitored through biweekly (online or in-person) debrief meetings with co-implementers and the research team. Interventions including video screening were adapted for the urban and rural contexts, with scheduling tailored to participant availability and appropriate language or dialect where needed. The co-implementers and intervention participants received reimbursement for transport costs.

### Data collection and analysis

Sociodemographic data were collected using a questionnaire covering age, sex, domicile, education, occupation, marital status, and clinical characteristics (type of leprosy, grade of disability, and type of LR). Participants reported LR type which was verified through clinician confirmation or review of medical records by healthcare workers when available. Acceptability of each intervention was assessed using a multi-faceted framework that captured the appropriateness of the interventions, based on participants’ perceptions. Following Sekhon et al.[42], seven concepts were used to evaluate acceptability: affective attitude (how participants felt about the intervention), burden, perceived effectiveness, ethicality, intervention coherence, opportunity costs, and self-efficacy (participants’ confidence in their ability to engage with and carry out the activities within the intervention). In this study, operational definition of seven acceptability concepts that guided the data analysis are described in Supplementary file 3.

Four types of participants were included:

a. Participants on treatment for LR who received one of the three interventions were assessed for knowledge, attitudes, and practices (KAP), mental well-being, and self-stigma at enrolment prior to initiation of the intervention and immediately after intervention completion. The assessments in Bahasa Indonesia (Supplementary file 4) were collected using Qualtrics and comprised: a validated KAP tool for measuring understanding of LR[43], the 14-point Warwick-Edinburgh Mental Wellbeing Scale²⁴, and two parts of SARI Stigma Scale (for disclosure concerns and internalized stigma)²⁵ (<u>Supplementary file 4</u>). The KAP scale comprised three domains. The knowledge domain scores ranged from 0–50, the attitude domain from 0–24, and the practice domain from 14–56. The knowledge domain scores were categorized as low (0–15), moderate (16–30), and high (31–50), while the attitude and practice domains were categorized using tertile-based cut-offs. Higher scores indicate greater knowledge, more favorable attitudes, and more appropriate practices. The Warwick-Edinburgh Mental Well-being Scale consists of 14 items scored on a 5-point Likert scale (1–5), yielding a total score ranging from 14 to 70. Higher scores indicate better mental well-being. The SARI Stigma Scale comprises four items of disclosure concerns and six items of internalized stigma scored on a 0–3 Likert scale, producing a total score ranging from 12-18 respectively. Higher scores indicate greater levels of stigma. In-depth interviews were conducted on completion of the intervention to explore the seven acceptability constructs and perceived changes in understanding of LR and self-care practices.
b. FM were interviewed immediately after watching the PV. The interviews explored their perceptions of the PV and perceived changes in their perspectives on providing physical and psychosocial support to individuals with LR who participated in the first group.
c. Co-implementers who implemented all three interventions were interviewed during the training week and again within two weeks of completion of the interventions. Pre- and post-intervention interviews explored the seven acceptability constructs, as well as co-implementers’ recommendations for improvement.
d. Healthcare professionals responsible for delivering LR services at the study sites were involved as non-participant observers of the interventions. For peer counselling and participatory video screening, they observed implementation from a distance without direct involvement. Meanwhile, telesupport group discussions were reviewed by the healthcare professionals retrospectively after the intervention had concluded. Twenty healthcare professionals from each study site took part in three focus group discussions (FGDs) within four weeks of completion of the interventions.

The FGDs with healthcare professionals in urban and rural settings facilitated reflection on key concepts related to the acceptability of the pilot interventions for the individuals with LR and the feasibility of integrating these peer-support models into existing leprosy care. Semi-structured guide for interviews and FGDs can be found in Supplementary file 2. All interviews and FGDs were audio-recorded, transcribed verbatim, and translated into English.

Data analysis employed a mixed-methods approach. Qualitative data were analyzed thematically in Atlas.ti. Descriptive statistics were used for KAP, WEMWBS, and SARI stigma data. All quantitative and qualitative data were anonymized before analysis by removing personal identifiers and assigning a unique participant code. Participant characteristics and outcomes were summarized by intervention group, setting (urban or rural), and time point (enrolment and post-intervention). Normality was assessed using Shapiro–Wilk tests and visual inspection of raw score boxplots. For non-normally distributed data, nonparametric methods were used. Within-group changes between baseline and post-intervention were analyzed using Wilcoxon signed-rank tests. Urban–rural differences at baseline and post-intervention were examined using Mann–Whitney U tests which stratified by intervention group. Differences across the three interventions were assessed using Kruskal–Wallis tests. All tests were two-tailed (p<0.05). Results are reported as medians with effect sizes calculated as r=Z/√N. Analyses were conducted in SPSS 27. Quantitative data were used to complement and triangulate qualitative findings.

### Ethics approval and consent to participate

This study obtained ethical approval from Dr. Soetomo General Academic Hospital (Approval ID: 0354/KEPK/I/2022). The interventions necessitated dialogue between Individual and counsellors on sensitive topics. We advocated for co-implementers to facilitate peer-counselling in a secure, private setting, agreed upon by both the counsellor and the beneficiary. All participants were informed about the study’s purpose, data confidentiality, voluntary participation, and their right to withdraw from the study at any point. After receiving comprehensive information, they signed a consent form in Bahasa Indonesia. Participants who were unable to read were provided with verbal information and recorded providing consent. No real or full names were identified in the transcripts or publications. All documents, photographs, and audio recordings were kept confidential and stored on a secure external hard drive, password-protected and accessible only to the research team. Personal data was handled in accordance with Indonesian law on information and electronic transactions (number 19/2016) and ministerial regulation (number 20/2016). Participants had the option to withdraw from the study and request the deletion of their data at any time.

## Results

### Roles and motivations of the co-implementers

All eight co-implementers who had completed treatment for LR acted as peer counsellors, facilitators of the telesupport group and participatory video makers. Four co-implementers were based in the urban setting and four in the rural setting. The co-implementers in the urban setting were three men and one woman. This diverse group included a retired man and three young adults. Three had completed senior high school, while one had not completed junior high school. In the rural setting, the co-implementers consisted of two men and two women who were grocery sellers, a student, and a housewife.

The co-implementers were actively involved in the intervention co-creation, adaptation, pilot implementation, and evaluation. In the baseline interviews, the co-implementers reported that their motivations for participating were to increase their understanding of LR and to act as a role model for peers. One said,

*“I want to know the impact of leprosy, because I have gone through it. What are symptoms of LR on others when they are treated [with corticosteroids], and how they feel. I joined to get better understanding and motivate others who are newly diagnosed.”* [S-003, Co-Implementer].

Another stated:

*“I want to tell them [individual with LR] that I am cured, a proof that cure is possible with the treatment.”* [B-003, Co-Implementer].

The co-implementers expressed their willingness to help provide better support to peers experiencing LR because they understood the challenges of living with the condition and recognized the importance of additional support, which they had not previously had. A co-implementer said, *“I participate to ensure the Madurese people continuing their [leprosy and LR] treatment in the community clinic. Then, [if they are treated] leprosy does not infect others”* [B-002, Co-Implementer]. Another explained, *“I was motivated to find out how I can support others with leprosy… So, we can be cooperative during the treatment and be healthy again together”* [B-001, Co-Implementer]

### Characteristics of intervention participants

Recruitment of participants took longer in community clinics because most individuals with LR were referred to and treated in the hospital. Recruitment for the three interventions required approximately two months in the hospital setting and five months in community clinics. A hundred participants received one of the three interventions. Sixty individuals had LR (22 (36.7%) in the urban setting) and 40 were family members (16 (40%) in the urban setting). Seven (11.7%) individuals with LR and one (2.5%) family member did not complete post-intervention assessments (see <u>Supplementary file 5</u>). Detailed information about the healthcare workers can be found in Supplementary file 5.

The sociodemographic characteristics of participants who received each intervention are presented in Table 1. The sex and ethnicity distribution were evenly balanced. One-third of participants were young adults (17-26 years old). More individuals with LR and their accompanying FM lived in rural areas. Approximately one-third were self-employed and almost one-quarter were unemployed. Fifty-four (54%) participants had completed high school. Most of the participants were Muslim and married. Forty (67%) individuals with LR had multibacillary leprosy and 44 of them had Grade-1 disability (73%). Thirty-six (60%) participants had T1R and 24 (40%) ENL.

**Table 1.** Sociodemographic and clinical characteristics of study participants by intervention.

| Characteristic | Individuals with Leprosy Reaction (n=60) |  |  | Family member | N (%) |
| --- | --- | --- | --- | --- | --- |
|  | Peer counselling (N= 25) | Telesupport group (N= 24) | Participatory Video (N= 11) | Participatory Video (N= 40) |  |
| **Sex** |  |  |  |  |  |
| Male | 26 | 36 | 9 | 15 | 53 (53%) |
| Female | 18 | 8 | 12 | 25 | 47 (47%) |
| **Age** |  |  |  |  |  |
| 18-26 | 8 | 13 | 2 | 8 | 31 (31%) |
| 27-36 | 8 | 5 | 3 | 12 | 28 (28%) |
| 37-46 | 4 | 4 | 2 | 9 | 19 (19%) |
| >46 | 5 | 2 | 4 | 11 | 22 (22%) |
| **Domicile area** |  |  |  |  |  |
| Urban | 8 | 8 | 6 | 16 | 38 (38%) |
| Rural | 17 | 16 | 5 | 24 | 62 (62%) |
| **Occupations** |  |  |  |  |  |
| Employee (full-time/part-time) | 3 | 9 | 4 | 5 | 21 (21%) |
| Self-employed (farmer, merchant) | 8 | 5 | 2 | 18 | 33 (33%) |
|  | 2 | 4 | 1 | 5 | 12 (12%) |
| Volunteers or students | 8 | 5 | 2 | 11 | 26 (26%) |
| Unemployed | 4 | 1 | 2 | 1 | 8 (8%) |
| Others |  |  |  |  |  |
| **Level of academic** |  |  |  |  |  |
| Unable to read and incomplete primary education | 1 | 1 | 1 | 1 | 4 (4%) |
| Literate and incomplete primary education | 0 | 1 | 1 | 5 | 7 (7%) |
| Completed primary school | 7 | 4 | 1 | 16 | 28 (28%) |
| Completed high school | 13 | 18 | 7 | 16 | 54 (54%) |
| University | 4 | 0 | 1 | 2 | 7 (7%) |
| **Ethnicity** |  |  |  |  |  |
| Javanese | 12 | 11 | 7 | 16 | 46 (46%) |
| Madurese | 12 | 12 | 4 | 24 | 52 (52%) |
| Others | 1 | 1 | 0 | 0 | 2 (2%) |
| **Religion** |  |  |  |  |  |
| Islam | 24 | 24 | 11 | 40 | 99 (99%) |
| Christian | 1 | 0 | 0 | 0 | 1 (1%) |
| **Marriage status** |  |  |  |  |  |
| Single | 9 | 12 | 2 | 11 | 34 (34%) |
| Married | 15 | 10 | 9 | 27 | 61 (61%) |
| Widowed | 1 | 1 | 0 | 2 | 4 (4%) |
| Separated | 0 | 1 | 0 | 0 | 1 (2%) |
| **WHO leprosy classification\*** |  |  |  |  |  |
| Multibacillary | 17 | 19 | 4 | N/A | 40 (67%) |
| Paucibacillary | 8 | 5 | 7 |  | 20 (33%) |
| **WHO Grade of disability\*** |  |  |  |  |  |
| Level 1 | 14 | 22 | 8 | N/A | 44 (73%) |
| Level 2 | 2 | 0 | 1 |  | 3 (5%) |
| No disability | 9 | 2 | 2 |  | 13 (22%) |
| **Type of Leprosy Reactions (LR)#\*** |  |  |  |  |  |
| T1R | 13 | 16 | 6 | N/A | 36 (60%) |
| ENL | 12 | 8 | 5 |  | 24 (40%) |
| *N/A: not applicable |  |  |  |  |  |
| #Participant-reported and clinically verified when available |  |  |  |  |  |

### Perceived acceptability of each intervention

The interventions were viewed as acceptable and beneficial by individuals with LR, healthcare workers, and co-implementers in both urban and rural settings. Table 2 presents quotes on the perceived acceptability of the three interventions. Interviewees from all groups recognized the value of peer involvement, however their perspectives differed. Individuals with lived experience of LR (participants on treatment for LR and co-implementers) emphasized empathy, belonging, and empowerment that led to improvement in LR participants’ motivation in adhering LR treatment during peer counselling and in the telesupport groups. FM emphasized improvement in their knowledge of leprosy and LR affecting the support they provided for their relative with LR. Meanwhile, healthcare workers reported improvements in communication and treatment adherence among participants with LR after participation in the interventions.

**Table 2.** Selected quotes of perceived acceptability among participants.

| Component | Peer Counselling | Telesupport group | Video Screening at healthcare facilities |
| --- | --- | --- | --- |
| Affective attitude | "Alhamdulillah, the activities were beneficial. I feel more motivated because [the co-implementer] encouraged me to recover. My husband was supportive when I talked about this [peer counselling] in puskesmas" [B101, individual with LR].<br>"Activities should be more frequent or more routine" [B103, individual with LR].<br>"Peer counselling was good. Several patients expressed their happiness to me. They made new friends. Their perception [of leprosy and its reactions] has changed. It [the counselling] has improved their self-esteem and excitement to visit the outpatient clinic." [FGD1S, Healthcare worker B].<br>"Enthusiastic again, knowing that this disease can be cured, so they gain new insight, who knows, maybe new ideas to broaden their horizons." [FGD1S, Healthcare worker D].<br>"It reduces the patient's anxiety about the [leprosy] reactions they experience. Peer counselling gives the most benefits" [FGD1B, Healthcare worker A]. | "I can get responses, input and suggestions from friends in the [telesupport group] group. If I tell friends or family, they might not understand" [S207, Individual with LR].<br>"I'm happy when I know another new patient [with LR]. We share experiences when we are ill" [S204, Individual with LR].<br>"Beneficial and provides a [telesupport group] platform where they can all be motivated to continue living better lives. Furthermore, it includes a group of people with leprosy reactions, which can be a place to exchange ideas" [FGD1B, Healthcare worker C]. | "I hope that I can gather with those people more often. Thank God, I'm happy, it means that this disease isn't just in me, it means a lot" [S305, Individual with LR].<br>"The video soothes me... Insha Allah, individual who experience leprosy [and LR] will be recovered" [S303, Individual with LR].<br>"I was not confident, but I feel better now." [S306, Individual with LR].<br>"It's also for our knowledge. For patients, it's okay, so don't feel inferior. You can show that video to provide information to our friends who experience it [leprosy reactions] so they won't hesitate to communicate." [S309, Individual with LR]. |
| Burden | "Sometimes, our conversations are interrupted because the nurse calls the peer counselee..." [S-002, Co-implementer]<br>"Another case was the patient was difficult to engage in a conversation due to stomach pain." [S-004, Co-implementer]<br>"After leaving the outpatient room, they have to go to the pharmacy, they don't go straight home or work. They need sparing more time for counselling..." [FGD1S, Healthcare worker A].<br>"There's no money for vehicle fuel [to visit community clinic]" [FGD1B, Healthcare worker A]. | "Sometimes we lacked communication... Some are less responsive." [S013, Co-Implementer]<br>"I hardly read the [WhatsApp] chat. I can't type, but my child assisted me" [B014, Co-implementer]<br>"As we have never met face to face..., we don't know if he doesn't tell us" [S014, Co-implementer]<br>"I rarely comment. I look at the chat, because I'm working, I don't have my phone with me." [S205, Individual with LR]<br>"Telesupport group is possible [to sustain], but some patients [with | "I hope it's [the duration] at most three minutes because not all people stay in that place for that long" [S413, Accompanying family member]<br>"This could be made more engaging in a shorter format." [FGD1S, Healthcare worker E].<br>"There's no TV or the [internet] connection is slow." [FGD2B, Healthcare worker C]<br>"Additionally, the video mentioned that they were cured, but they didn't show any signs of the symptoms they experienced beforehand. With the before and after, people who saw it will |
|  | "The distance between the patient's home and the community clinic is too far..." [FGD1B, Healthcare worker A, B, D & F].<br>"Peer counselling requires funding from the community clinic for remuneration" [FGD1B, Healthcare worker C].<br>"We don't have [resources and capacity for] training" [FGD1S, Healthcare worker A]. | LR] are elderly and some can't access social media. Signal and internet package issues too." [FGD1B, Healthcare worker A]. | believe it more." [FGD1B, Healthcare worker E]. |
| Ethicality | "We, as healthcare workers, or perhaps their families, cannot empathize with their condition. We don't experience it [LR] ourselves, unlike people affected by leprosy. From a supportive perspective, it helps psychologically." [FGD1S, Healthcare worker B].<br>"There is a sense of shared destiny and struggle. This [peer counselling] allows them to better understand each other when communicating..." [FGD2B, Healthcare worker C].<br>"Patients [with LR] gain additional information from their peers, thus alleviating burden on healthcare workers." [FGD2B, Healthcare worker E]. | "Helping others affected by leprosy and increasing knowledge, so we don't get stressed, and our reactions don't get worse." [S203, Individual with LR]<br>"With that [WhatsApp] group, it's easier to share information." [FGD1B, Healthcare worker E].<br>"Yes, the information [about LR] is getting more widely available via that telesupport group." [FGD2B, Healthcare worker A,B,C,D & E].<br>"These co-implementers are not medical professional, so there is a possibility of distortion of information." [FGD1S, Healthcare worker C]. | "I motivate those who are sick to be cheerful again, to be healthy again." [S411, Accompanying Family Member].<br>"I must support to provide knowledge to those with leprosy who should believe they can be cured." [B423, Accompanying Family Member].<br>"[This video] helps people who don't know about leprosy [to understand about LR]." [FGD2B, Healthcare worker A].<br>"Sometimes people's feelings are different. ... I'm afraid others [with LR] are embarrassed" [S411, Accompanying Family Member]. |
| Intervention coherence | "There is a place to complain or talk about any matters with someone who has previously had this disease. So, I feel like I am not alone in this pain" [S104, Individual with LR]<br>"[Individual with LR] said, 'I don't have enough friends. I can't talk to anyone when I am at work or at home. I can't even express my feelings to the neighbors. Here, I can just chat with you.'" [S011, Co-implementer]<br>"Peers are very supportive for patients with leprosy [reactions]." [FGD2B, Healthcare worker C].<br>"They're more communicative and care about their disease. So, they help healthcare workers in | "I often talk about emotional control. I share how to control emotions, how to calm yourself." [S210, Individual with LR]<br>"We get motivation that this disease [LR] can be cured." [S205, Individual with LR]<br>"Telesupport group allows sharing information among patients [with LR] and updating each other's condition." [FGD1B, Healthcare worker A & D].<br>"The WhatsApp group helps them [individuals with LR] to obtain guidance from their peer" [FGD1S, Healthcare worker D].<br>"Maybe, they'll update questions they're unsure about to us, so they can ask it directly to prevent | "The video tells them that there are people who can recover" [FGD1S, Healthcare worker].<br>"[Through this video] This is how you can prevent others to stay away from people with the disease" [S405, Accompanying family member]<br>"The people in the video tell stories about their recovery process... I want to be like everyone else in the video, healthy." [B301, Individual with LR]<br>"I remember earlier, 'I have leprosy, it's not hereditary, it's bacterial'...." [S303, Individual with LR]<br>"I feel calm when watching the Madurese man who told me [the |
|  | *providing information about leprosy [and LR]." [FGD2B, Healthcare worker B].* | *misinformation."* [FGD1S, Healthcare worker C]. | *audience] to challenge yourself..." [S303, Individual with LR]*<br>*"The symptoms are different for everyone... The video gives many examples for me and for everyone." [S305, Individual with LR]*<br>*"It would be better if it's shown on a larger screen so that it could be seen by the wider community out there" [S411, Accompanying family member]* |
| Opportunity costs | *"Finding people who volunteer to join the [peer-counselling] team and put aside all their work isn't that easy, right? Working without pay is definitely impossible." [FGD1S, Healthcare worker D].* | *"If someone asks a question that the co-implementers didn't understand, the health worker can help explain it to us" [S203, Individual with LR].*<br>*"Maybe if healthcare workers join the [WhatsApp] group, they [individuals with LR] won't freely share their stories." [FGD1B, Healthcare worker E].* | *"Adding the before-and-after condition, like when you first got the disease, the symptoms... Most people with this kind of disease are afraid they won't be able to go back to their original state" [S307, Individual with LR]* |
| Perceived effectiveness | *"If I have a complaint, I can tell stories to my peers. In the past, I delayed for 1 month or 2 weeks. I'm always late. Now, thank God, I'm starting to change that bad habit, and my enthusiasm is starting to increase" [B106, Individual with LR]*<br>*"I am able to clear my mind after discussing with [the co-implementer]. So, I don't get tired, don't overthink, and the [leprosy] reactions don't recur." [S105, Individual with LR]*<br>*"I was not allowed to participate in family gatherings; now my family has allowed me to" [B014, Co-implementer]*<br>*"We, as medical personnel, only focus on treatment... An external support system is important to help or at least to alleviate their stress" [FGD1S, Healthcare worker B].*<br>*"The patients [with LR] come to us hoping for a cure, but that [leprosy] reaction makes them depressed. With peer counselling, they understand that this may happen to some* | *"They [co-implementers] are always supportive. Occasionally, I'm late for treatment, and they remind me that it's time for treatment" [S206, Individual with LR]*<br>*"Once I lost [to follow up] regular patient, accidentally [a co-implementer] reported to me 'Ma'am, [an individual with LR] passed away'... They [the co-implementer] said the information was shared in that group... So, it's easy for me to report it to the Health Office." [FGD1S, Healthcare worker A].*<br>*"The [individuals with LR] become more diligent in their checkups" [FGD1S, Healthcare worker A].*<br>*"From that [telesupport group] group, patients learn about leprosy." [FGD1B, Healthcare worker C].*<br>*"They [individuals with LR] seem to be motivated to join a larger forum..." [FGD1S, Healthcare worker A].* | *"Well, if you watch that video, why should I be embarrassed?" [S305, Individual with LR]*<br>*"Since watching that video, I've been more regular with my treatment" [B320, Individual with LR]*<br>*"The video shows that young people, old man, and even someone at my age [affected by leprosy reactions].... It can affect anyone, regardless of age. It doesn't discriminate between ages. It's not hereditary." [S305, Individual with LR]*<br>*"It's ocular proof because this video is told directly by a patient who experienced leprosy reactions until he was cured. So that others believe it." [B407, Accompanying family member]*<br>*"Now, I believe that leprosy can be cured." [B417, Accompanying family member]*<br>*"It uses simple language that's not too overly complex seems to be acceptable to all groups" [FGD1S, Healthcare worker A].*<br>*"The patient [with LR] has better acceptance due to the video* |
|  | people, and it can be treated... They have a better acceptance" [FGD1S, Healthcare worker A]<br>"Some patients become calmer." [FGD1B, Healthcare worker C].<br>"The success is reassuring them [individual with LR] that they're not alone" [S012, Co-implementer]<br>"The peer counselling feels more familial, more person-to-person, more like a friend. Third, the person feels less inferior." [FGD1S, Healthcare worker A].<br>"There's a change... he takes the initiative to seek treatment himself." [FGD1B, Healthcare worker B] |  | instead of listening to the healthcare worker's explanation. This video helps us in educating patients" [FGD2B, Healthcare worker]. |
| Self-efficacy | "The counsellor and I have almost the same thoughts, there's no need to be discouraged by a disease like this..." [S106, Individual with LR]<br>"I wish the counselees were the same age as me so I could respond to them better." [S013, Co-implementer]<br>"We became more enthusiastic and had friends" [B011, Co-implementer]<br>"Yesterday, I communicated with him about his check-up [and our counselling session]." [S013, Co-implementer]<br>"Because they share the same fate, experiencing leprosy..." [FGD2B, Healthcare worker E]<br>"If possible, all community clinics should have peer counselling. So that everyone can get the benefit. Maybe the [counselling] days can be adjusted to the patient's outpatient visit schedule." [FGD1B, Healthcare worker E] | "We just want to convey something, but I'm afraid of being misunderstood" [S209, Individual with LR].<br>"I'm ready to be a peer supporter" [B210, Individual with LR].<br>"I can tell my concerns [in the group]." [S210, Individual with LR]<br>"Sharing with friends the joys and sorrows of living or experiencing leprosy. Receiving feedback from friends" [S204, Individual with LR]<br>"We casually talk and make a joke about our experience, it's good, so it can be more relaxed." [S013, Co-implementer]<br>"I always get cut off [by other co-implementers], It could make the participants confused" [S012, Co-implementer] | "I'm motivated by someone who had leprosy and can return to their healthy body." [S307, Individual with LR]<br>"If I have a concept [of the video], I'll be confident [to be one of the video makers]." [S308, Individual with LR]<br>"The family seems to be more supportive after watching the video because they're being educated and informed." [FGD1S, Healthcare worker A] |

Individuals with LR reported finding the interventions coherent, beneficial, and effective in the interviews. They felt more confident in their ability to manage LR after the interventions. Both individuals with LR and family members said they were confident to participate in the intervention. These opinions frequently emerged in FGDs with healthcare workers.

#### Affective attitude

Emotional responses among participants with LR, co-implementers and healthcare workers varied across the three interventions. Across settings, individuals with LR viewed peer counselling interactions as motivating and emotionally supportive. They described counselling sessions as comforting and trustworthy (Table 2), and this was echoed by the healthcare workers in FGDs. In the interviews, most participants with LR in both settings felt the benefits of sharing their concerns with and asking questions of co-implementers. This was reported by urban-based healthcare workers. In the urban setting, peer counselling was deemed as a positive experience which renewed enthusiasm and increased self-esteem by LR participants and healthcare workers. Co-implementers suggested incorporating peer counselling into routine clinic activities. Two individuals with LR from the rural setting suggested frequent counselling sessions including family members. The rural healthcare workers stated that peer counselling reduced affected persons’ anxiety associated with LR. Retention was high (see <u>Supplementary file 5</u>) with 21 out of 25 (84%) individuals with LR completing five counselling sessions and the post-intervention assessment.

Individuals with LR in the telesupport groups appreciated the sense of connection between the members of the group. In urban and rural groups, they perceived telesupport provided a flexible platform for sharing information and experiences about LR and expressing their thoughts. In the interviews, individuals with LR described feelings of happiness in making new friends virtually and emotional reassurance derived from connecting with co-implementers and others with similar experiences of LR. The telesupport group was perceived by individuals with LR as a valued space for mutual encouragement and exchange of practical advice. Healthcare workers had similar perceptions highlighting telesupport role in fostering motivation for improved health and collective problem-solving.

The participatory video was perceived as a feasible and optimistic by showing recovery is possible in both settings (Table 2). In the post-intervention interview, individuals with LR and accompanying FM described that the video helped viewers recognize that leprosy and LR are treatable, non-hereditary conditions with diverse presentations. They said that personal recovery narratives in the video encouraged aspirations of recovery for individuals with LR. The individuals with LR mentioned that the video helped them believe that LR could be treated successfully and that they were not alone in facing the condition. Healthcare workers in both settings had a positive response to the video.

#### Burden

In peer counselling, individuals with LR in the urban setting experienced scheduling challenges which were echoed by the co-implementers. The co-implementers mentioned that counselling sessions were often interrupted by patients’ clinical assessments. LR participants in the rural setting found it challenging to routinely attend counselling sessions at the community clinic due to distance and associated high transportation cost. In the FGD, healthcare workers in rural settings said that long travel distances hindered LR participants from attending sessions on time. They explained that community clinics lacked sufficient funding to cover transportation costs if peer counselling were integrated into routine services in their healthcare facility.

Telesupport groups were perceived as a challenge by four individuals who were unable to read or had limited spare time. Those unable to read said they only participated using audio messages or were helped by their children. They admitted being reluctant to initiate chats and one participant was reluctant to express her feelings and respond to questions from friends in the WhatsApp group because she was afraid of giving the wrong answer, which could lead to misunderstandings (Table 2). Some individuals were occasionally unable to be active in the WhatsApp group and respond to the group discussion in real time due to work commitments. Co-implementers noted that emotional bonding could be harder to establish and sustain without face-to-face meetings. Structural barriers—such as older age, limited familiarity with the messaging application, unstable internet connectivity, and the cost of mobile data packages—were highlighted by healthcare workers as challenges to sustained participation in rural.

Three individuals with LR in the urban setting would have preferred a bigger device screen for viewing the participatory video. Another two requested descriptions of illness trajectory of the co-implementers in the PV from leprosy diagnosis and onset of LR to recovery. Healthcare workers also thought this was important as “before and after” symptom description could be more compelling than personal narratives after recovery. The healthcare workers and accompanying FM suggested that future videos should be shorter to a 3-minute video to maintain viewer attentiveness particularly where time constraints within healthcare facilities limited participants’ ability to engage with longer videos. Healthcare workers emphasized that some clinics lacked televisions or had unstable internet connections which made the video screening less feasible when streaming was required. Most individuals with LR suggested uploading the video onto publicly available platforms, but healthcare workers and accompanying family members preferred to show it only in supportive environments where viewers could engage with the content privately and without fear of being stigmatized.

#### Ethicality

The three peer support interventions were regarded by all participants and healthcare workers as ethically appropriate and well aligned with the needs of individuals with LR. In peer counselling, LR participants said they could share their concerns or ask co-implementers questions. The healthcare workers believed that counselling sessions could alleviate their burden in providing education about LR and that information exchange between peers provided a psychologically supportive and complementary component of care. Healthcare workers emphasized that clear guidance and referral pathways were needed for clinical questions raised during peer counselling and telesupport activities.

Individuals with LR deemed the telesupport group reduced stress and concerns about worsening LR through shared information and emotional support. Healthcare workers recognized the value of telesupport in facilitating wider access to information and peer-to-peer knowledge exchange. However, ethical concerns were raised by healthcare workers in the urban setting regarding the potential for misinformation in counselling sessions and telesupport groups. The healthcare workers highlighted the need for oversight and training to ensure the accuracy of information shared.

Individuals with LR and accompanying FM viewed video screening as a means of motivating affected individuals to regain confidence and believe in the possibility of recovery. However, ethical concerns were raised by FM and individuals with LR regarding potential emotional discomfort. Participants acknowledged that affected individuals might feel embarrassed or exposed to public screening of the video.

#### Intervention coherence

Individuals with LR across settings demonstrated clear understanding of the purpose and function of the interventions. They described peer counselling sessions in the interviews as a safe space to discuss fears, stigma, and daily challenges that were often difficult to share with healthcare workers. Many individuals with LR reported feeling a sense of community after the sessions. The healthcare workers observed that the co-implementers were communicative in providing information about LR complementing healthcare workers’ efforts in both settings.

The LR participants and co-implementers in interviews described telesupport group as a platform for sharing information, providing peer guidance, and supporting emotional regulation. The telesupport group was perceived by individuals with LR as a space to update others on health status and seek clarification on uncertainties for reducing misinformation.

LR participants in the interviews understood the video’s purpose and retained key messages about medication adherence and self-care. Several individuals with LR and family members reported a shift in understanding from believing leprosy to be a hereditary disease and recognizing it as caused by a bacterial infection. In the FGD, healthcare workers perceived the video as a practical and engaging education tool to improve conversations about LR and reduce fear among newly diagnosed patients.

#### Opportunity costs

In both settings, individuals with LR stated counselling sessions utilized time which had to be spent in the clinical facility because they were conducted whilst waiting for assessments or medication. However, peer counselling was perceived by healthcare workers and co-implementers as involving significant opportunity costs because of the time demands and potential loss of income.

Individuals with LR and healthcare workers felt the opportunity costs of the telesupport group were primarily associated with trade-offs between professional oversight and participants’ willingness to engage openly. The involvement of healthcare workers was perceived by individuals with LR as beneficial for clarifying complex or uncertain information. Healthcare workers viewed their presence as potentially limiting the affected individuals’ freedom to share personal experiences.

The 9-minute-long PV screening was not perceived as a significant burden by participants with LR because it occurred whilst waiting at the health facility. Some family members reported that the video slightly extended their time at the clinic, but they considered the information and reassurance gained outweighed the additional time required. Healthcare workers reported the video screening required coordination and occasional assistance from the research team or co-implementers. They explained that if adopted into routine practice, the video would require dedicated devices and stable electricity supply to show the video.

#### Perceived effectiveness

Peer counselling was perceived by individuals with LR as effective in improving psychological well-being, coping, and care-seeking behaviors. In the hospital setting, individuals with LR stated participation in peer counselling helped with managing and preventing LR recurrence by recognizing its triggers, reducing stress, and improving self-care practices. Rural participants mentioned that counselling increased their understanding of LR by the third counselling session and enabled them to have more informed discussion with family members. Individuals with LR said that hearing recovery stories from co-implementers increased their confidence and normalized the experience of living with LR. Healthcare workers in FGDs believed peer counselling increased understanding and acceptance of LR among the affected individuals. The healthcare workers in the urban setting described that most LR affected participants in the urban setting were more confident asking questions during clinical assessment, while healthcare workers in rural setting reported LR affected participants attended appointments more regularly. Healthcare workers reported that individuals who received peer counselling were more determined to adhere to treatment for LR, in rural settings appeared to reduce anxiety associated with LR and improve understanding of LR, self-care practice, and treatment.

The telesupport group was perceived as effective in supporting treatment adherence. LR participants from urban setting were more active in the discussion forums than rural participants. During the observation period, there was a median of 268 messages per week in the urban group compared to a median of 43 messages per week in the rural group. Individuals with LR said they learned about medication, side effects, and self-care through the telesupport group. The telesupport participants reportedly felt calmer because the co-implementers provided encouragement if a participant experienced difficulties coping with LR. The participants of the telesupport group expressed greater confidence and less feelings of loneliness in facing their LR. All group participants felt happy to be able to share their concerns with other group members (Table 2). Four healthcare workers reported changes in how individuals with LR communicated with them after participating in the telesupport group. They described individuals as more confident in asking questions and increased knowledge. Two individuals with LR described receiving timely reminders to maintain regular treatment for LR. Healthcare workers highlighted the group’s role in facilitating communication about patient status, in particular for identifying individuals lost to follow-up and even death through group updates shared by the participants. Healthcare workers believed that the group reduced their workload for minor queries by enabling participants to seek support directly from co-implementers, although this reduction was not formally measured. The approach was perceived as low cost because it relied on existing mobile messaging platforms and did not require additional infrastructure or resources to maintain communication and motivation.

The PV was perceived as effective by all intervention participants and healthcare workers because it used first-person recovery narratives. Family members described the videos as providing credible and “visual” evidence that leprosy and LR can be cured. Healthcare workers said that the use of simple language and diverse representations of age reinforced clear messages and was perceived to motivate improved treatment adherence.

#### Self-efficacy

The involvement of people with past experience of LR as co-implementers was perceived by individuals with LR and healthcare workers as being relatable. Both co-implementers and individuals with LR gained confidence in communicating about LR and sharing experiences. The urban individuals with LR explained that they had similar experiences with the co-implementers and gained better understanding about the condition. The individuals with LR in the rural setting felt more comfortable after completing the counselling session. Most participants felt that peer counselling should become a routine component of LR services, although LR-affected participants doubted their abilities to support their peers, the co-implementers felt confident and proud of their role. Healthcare workers in both settings said that shared lived experience fostered mutual encouragement.

The telesupport group elicited mixed impacts on self-efficacy among participants and co-implementers. Some individuals with LR described increased confidence in expressing concerns and supporting others. In the interviews, they expressed willingness to take on peer-support roles in telesupport groups. However, one individual with LR reported fears of being misunderstood or disrupting group conversations.

The PV was perceived to enhance self-efficacy among individuals with LR and their family members. Individuals with LR described feeling motivated by observing peers and strengthened belief in their own capacity to improve health outcomes. Some individuals with LR also expressed readiness to contribute actively to future video development. For many, visual storytelling and participation of the co-implementers affected by LR made complex information more relatable and gave authenticity to the content. In the interviews, accompanying FM expressed greater support for the individuals with LR following viewing of the video. Despite this, some individuals with LR said that they remained reluctant to disclose their condition to their extended family due to persistent stigma. Healthcare workers perceived that if video screening increased family understanding, this could foster more supportive environments and reinforce participants’ confidence.

### Changes in scores for knowledge, attitude, practice, mental well-being, and self-stigma following intervention

The median knowledge score of each group of urban participants was higher than those of the corresponding group in the rural settings prior to the interventions, but these differences were not statistically significant for any interventions (Table 3). The knowledge scores of urban participants improved significantly following telesupport (P=0.01) and PV screening (p=0.04). There was no significant change in the knowledge scores of urban participants who received peer counselling. The knowledge scores of rural participants significantly improved following Telesupport (P=0.01). Pre-intervention attitude scores (<u>Supplementary file 6</u>) were significantly higher in urban participants compared to rural participants in the peer counselling (p=0.002) and telesupport groups (p<0.001). Attitude scores increased significantly following peer counselling in both urban (p=0.026) and rural participants (p=0.042) (Table 3). Practice related to LR management scores (<u>Supplementary file 6</u>) before the interventions did not differ significantly between urban and rural groups for any intervention and no statistically significant changes in scores were observed following the interventions.

**Table 3.**
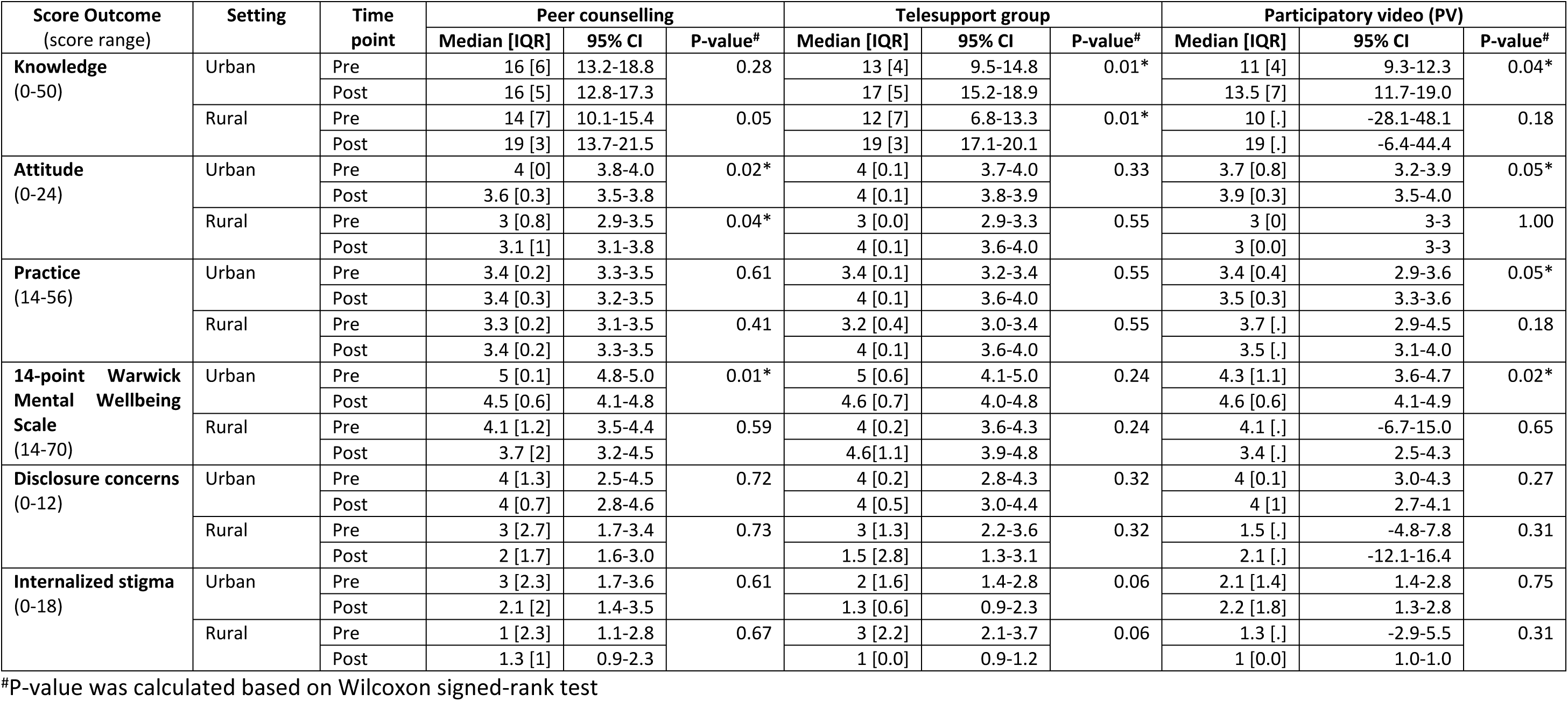
Summary of score outcomes in knowledge, attitude, practice, mental well-being, and self-stigmass.

| Score Outcome<br>(score range) | Setting | Time<br>point | Peer counselling |  |  | Telesupport group |  |  | Participatory video (PV) |  |  |
| --- | --- | --- | --- | --- | --- | --- | --- | --- | --- | --- | --- |
|  |  |  | Median [IQR] | 95% CI | P-value<sup>#</sup> | Median [IQR] | 95% CI | P-value<sup>#</sup> | Median [IQR] | 95% CI | P-value<sup>#</sup> |
| **Knowledge**<br>(0-50) | Urban | Pre | 16 [6] | 13.2-18.8 | 0.28 | 13 [4] | 9.5-14.8 | 0.01* | 11 [4] | 9.3-12.3 | 0.04* |
|  |  | Post | 16 [5] | 12.8-17.3 |  | 17 [5] | 15.2-18.9 |  | 13.5 [7] | 11.7-19.0 |  |
|  | Rural | Pre | 14 [7] | 10.1-15.4 | 0.05 | 12 [7] | 6.8-13.3 | 0.01* | 10 [.] | -28.1-48.1 | 0.18 |
|  |  | Post | 19 [3] | 13.7-21.5 |  | 19 [3] | 17.1-20.1 |  | 19 [.] | -6.4-44.4 |  |
| **Attitude**<br>(0-24) | Urban | Pre | 4 [0] | 3.8-4.0 | 0.02* | 4 [0.1] | 3.7-4.0 | 0.33 | 3.7 [0.8] | 3.2-3.9 | 0.05* |
|  |  | Post | 3.6 [0.3] | 3.5-3.8 |  | 4 [0.1] | 3.8-3.9 |  | 3.9 [0.3] | 3.5-4.0 |  |
|  | Rural | Pre | 3 [0.8] | 2.9-3.5 | 0.04* | 3 [0.0] | 2.9-3.3 | 0.55 | 3 [0] | 3-3 | 1.00 |
|  |  | Post | 3.1 [1] | 3.1-3.8 |  | 4 [0.1] | 3.6-4.0 |  | 3 [0.0] | 3-3 |  |
| **Practice**<br>(14-56) | Urban | Pre | 3.4 [0.2] | 3.3-3.5 | 0.61 | 3.4 [0.1] | 3.2-3.4 | 0.55 | 3.4 [0.4] | 2.9-3.6 | 0.05* |
|  |  | Post | 3.4 [0.3] | 3.2-3.5 |  | 4 [0.1] | 3.6-4.0 |  | 3.5 [0.3] | 3.3-3.6 |  |
|  | Rural | Pre | 3.3 [0.2] | 3.1-3.5 | 0.41 | 3.2 [0.4] | 3.0-3.4 | 0.55 | 3.7 [.] | 2.9-4.5 | 0.18 |
|  |  | Post | 3.4 [0.2] | 3.3-3.5 |  | 4 [0.1] | 3.6-4.0 |  | 3.5 [.] | 3.1-4.0 |  |
| **14-point Warwick**<br>**Mental Wellbeing**<br>**Scale**<br>(14-70) | Urban | Pre | 5 [0.1] | 4.8-5.0 | 0.01* | 5 [0.6] | 4.1-5.0 | 0.24 | 4.3 [1.1] | 3.6-4.7 | 0.02* |
|  |  | Post | 4.5 [0.6] | 4.1-4.8 |  | 4.6 [0.7] | 4.0-4.8 |  | 4.6 [0.6] | 4.1-4.9 |  |
|  | Rural | Pre | 4.1 [1.2] | 3.5-4.4 | 0.59 | 4 [0.2] | 3.6-4.3 | 0.24 | 4.1 [.] | -6.7-15.0 | 0.65 |
|  |  | Post | 3.7 [2] | 3.2-4.5 |  | 4.6 [1.1] | 3.9-4.8 |  | 3.4 [.] | 2.5-4.3 |  |
| **Disclosure concerns**<br>(0-12) | Urban | Pre | 4 [1.3] | 2.5-4.5 | 0.72 | 4 [0.2] | 2.8-4.3 | 0.32 | 4 [0.1] | 3.0-4.3 | 0.27 |
|  |  | Post | 4 [0.7] | 2.8-4.6 |  | 4 [0.5] | 3.0-4.4 |  | 4 [1] | 2.7-4.1 |  |
|  | Rural | Pre | 3 [2.7] | 1.7-3.4 | 0.73 | 3 [1.3] | 2.2-3.6 | 0.32 | 1.5 [.] | -4.8-7.8 | 0.31 |
|  |  | Post | 2 [1.7] | 1.6-3.0 |  | 1.5 [2.8] | 1.3-3.1 |  | 2.1 [.] | -12.1-16.4 |  |
| **Internalized stigma**<br>(0-18) | Urban | Pre | 3 [2.3] | 1.7-3.6 | 0.61 | 2 [1.6] | 1.4-2.8 | 0.06 | 2.1 [1.4] | 1.4-2.8 | 0.75 |
|  |  | Post | 2.1 [2] | 1.4-3.5 |  | 1.3 [0.6] | 0.9-2.3 |  | 2.2 [1.8] | 1.3-2.8 |  |
|  | Rural | Pre | 1 [2.3] | 1.1-2.8 | 0.67 | 3 [2.2] | 2.1-3.7 | 0.06 | 1.3 [.] | -2.9-5.5 | 0.31 |
|  |  | Post | 1.3 [1] | 0.9-2.3 |  | 1 [0.0] | 0.9-1.2 |  | 1 [0.0] | 1.0-1.0 |  |

<sup>#</sup>P-value was calculated based on Wilcoxon signed-rank test

The Warwick-Edinburgh Mental Wellbeing Scale scores (Table 3) were significantly lower in the rural participants compared to their urban counterparts in the peer counselling (p<0.001) and telesupport groups (p=0.007) prior to the intervention. Only the members of the urban PV group showed a statistically significant improvement in their 14-point Warwick-Edinburgh Mental Wellbeing Scale scores (p=0.028).

Self-disclosure scores (Table 3) were significantly lower in rural participants in the PV group compared to their urban counterparts (p=0.039) prior to the intervention. There were no significant changes in scores following the interventions. Internalized stigma scores (Table 3) were significantly higher in urban participants in the telesupport group compared to rural participants (p=0.011). There were no significant changes in internalized stigma scores following the interventions. Overall, quantitative findings aligned with qualitative data, indicating that intervention effects were shaped by both intervention modality and contextual setting.

## Discussion

To our knowledge, this is the first study to co-create and pilot three peer support interventions with individuals with LR in healthcare settings. This study contributes to the growing evidence base on holistic approaches to leprosy care and offers practical insights for scaling up community-driven interventions in leprosy-endemic areas. The findings provide new evidence on how different peer support interventions can be adapted to real-world health systems across different settings. Qualitative findings revealed why the interventions were acceptable, while quantitative results provided supportive signals of change in knowledge, attitude, practices, wellbeing, and self-stigma scores consistent with participant narratives. Across both sites, the interventions were well accepted, as shown by high participation and completion rates.

This mixed-methods study suggests that each intervention influences psychosocial and behavioural outcomes in a different way, shaped by contextual factors. Participants’ narratives revealed that acceptability stemmed from shared experience, trust, and relevance to the individual’s daily life. Emotional safety and mutual understanding were perceived as emerging themes on participants’ motivation to take part in the interventions. Within Sekhon’s framework of acceptability[42], intervention coherence, perceived effectiveness, and self-efficacy frequently emerged. The quantitative findings showed that individuals with LR in peer counselling group had an improvement in knowledge, attitude, and practices score in rural settings which were consistent with qualitative accounts emphasizing peer counselling as a safe space for empathy and emotional reassurance. Telesupport was primarily associated with improved knowledge in both settings supporting its role as an accessible information-delivery mechanism. Video screening demonstrated improvements in knowledge among urban and rural participants. Digital and visual tools have been reported to fill critical gaps where access to health information and clinical support are limited because of road infrastructure, limited public transportation availability, and expensive travel cost[50]. This pattern aligns with qualitative findings suggesting that this single episode intervention was more effective for increasing understanding of LR than the other two interventions with multiple interactions. There were substantial urban–rural differences prior to administration of all three intervention, particularly for attitude and mental well-being, which underscores the importance of accounting for contextual variation when interpreting intervention effects and planning scale-up. Together, these findings highlight the need for context-sensitive combinations of intervention strategies rather than a one-size-fits-all approach, to address the psychosocial dimensions of leprosy and LR.

A key identified characteristic of peer counselling was the motivational aspect across both settings. Qualitative findings emphasize empathy, shared lived experience, and trust as key mechanisms to explain the changes which were observed in knowledge and attitude across settings. Counselling sessions in this study offered a safe space to discuss fear, shame, and uncertainty. The counselling sessions helped to fill in the communication gap between LR-affected individuals and healthcare workers. Interactions with healthcare workers are often hampered by time constraints, hierarchical relationships, and a focus on biomedical outcomes. Because of these constraints, individuals with LR may feel less comfortable sharing sensitive or stigmatized experiences. Peer counselling also allowed medical information to be interpreted through lived experience of LR and making the discussion personally relevant and actionable. This conversation helped motivate individuals to adhere with treatment and strengthened self-efficacy. These mechanisms align with evidence from leprosy, tuberculosis, and HIV[19,44,45], where experiential authority and peer empathy are central drivers of engagement and behavior change. However, this study found that peer counselling required the greatest logistical coordination and opportunity costs, particularly for volunteer co-implementers, underscoring the importance of supervision, scheduling integration, and remuneration for sustainability.

The telesupport group and PV interventions built upon similar principles to peer counselling but differed in how they created connection, trust and learning. Peer counselling established trust through face-to-face interaction and shared lived experience, while telesupport and PV created engagement through interactive digital communication and storytelling. The telesupport group in this study functioned primarily as a rapid exchange of advice and reminders about outpatient visits. The telesupport group was valued in rural clinics, where distance and transport costs make access to in-person care more challenging. These findings explain the large knowledge gains observed in rural sites where access to information about LR was more limited than urban settings. The study findings echo evidence from digital peer support interventions for chronic diseases in low-middle income countries[25], which show that telesupport groups can improve knowledge and continuity of care, but risks of excluding the most marginalized members of the population[26,48,49]. Participation in telesupport groups depends on digital literacy and access to devices and connectivity, which are often less available to individuals who have experienced socioeconomic or social disadvantages.

The PV worked as both an educational tool to humanize LR and create dialogue within families. Watching peers share stories of recovery and coping created emotional resonance and initiated discussion between affected individuals and their relatives. The visual format was accessible to participants regardless of literacy levels and contributed to ethicality and perceived effectiveness as two key components of acceptability. The intervention was particularly effective in urban settings where significant improvements in knowledge, attitude, and practice were observed. In rural settings, the improvement was observed in mental well-being score. Adapting content to shorten the video to maintain viewer attention, was suggested as a potential improvement.

In the urban hospital setting, peer counselling played a more prominent role in facilitating emotionally sensitive conversations that digital platforms could not replicate. In rural clinics, where access to information and peer connection was limited, telesupport and video interventions addressed critical informational and social isolation needs. These findings emphasize that peer-support interventions thrive when aligned with existing resources, communication norms, and health-system structures.

Participants with LR, accompanying family members, co-implementers, and healthcare workers in this study consistently described each intervention as complementary to the standard LR care. Peer involvement bridged the clinical and lived experience of LR which further enhanced understanding, communication and trust between healthcare providers and patients. Collaboration between peers, affected individuals and healthcare workers reinforced mutual respect and shared responsibility in LR care as peer-based empathy fosters engagement, self-management, and social participation in chronic and stigmatized conditions such as HIV and mental illness[19,51], where peer-support interventions for LR can enhance routine clinical practice when appropriately integrated.

The findings suggest that the peer support interventions could be combined and adapted based on individual needs and local circumstances. We support selective scale-up of the interventions and context-sensitive integration. Each intervention fits a particular health system niche. Peer counselling provides unique emotional depth, trust, and knowledge but requires dedicated space, time, supervision, and counsellor remuneration to remain sustainable. Integrating peer counselling into routine care may reduce additional travel but scheduling separate sessions could allow more focused discussions. The individual circumstances of those utilizing the service may dictate the best model.

A telesupport group is most feasible where connectivity and digital literacy are sufficient. In areas with limited internet connection, low-tech or hybrid models, such as facilitator-led voice groups or scheduled messaging, may maintain inclusion or introduce a potential risk of misuse. The telesupport participants may be exposed to misinformation, breaches of confidentiality, or inappropriate interactions —including bullying or stigmatizing comments—particularly in contexts where leprosy remains highly sensitive and real-time facilitation is limited. To mitigate such harms, active moderation is essential, including clear group guidelines, trained facilitators, and mechanisms to address inappropriate behavior promptly.

Short locally produced participatory videos are a scalable awareness and education tool, well suited for healthcare and community settings. Videos accompanied by facilitated discussion to strengthen reflection and avoid passive viewing may be more effective but require additional resources. However, any video dissemination should be careful to minimize harms of those viewing and appearing in the video and be aware of the potential for the disclosure of their leprosy diagnosis to the wider community and for stigmatization. A balanced strategy may be needed such as combining flexible formats (e.g., shorter and shareable videos), moderated platforms, or facilitated video viewing in supportive environments.

Infrastructure, human resource capacity, and sociocultural context must guide decision-making rather than standardizing interventions across settings. This mirrors WHO recommendations for person-centered NTD management[52], which advocate modular peer-support interventions built around local capacity. Peer connection, regardless of intervention modality, transforms information on LR into meaning. The study participants with LR reported that peers lived experience made guidance trustworthy and relatable, and of peer co-implementers described greater confidence in supporting others. Healthcare workers recognized that peer-led communication reached patients in ways clinical advice could not. This triangulation reinforces that emotional safety and shared identity are fundamental enablers of acceptability. For future integration of peer support interventions in routine care, important operational factors include training and support, financial investment, technical support for digital interventions and culturally appropriate materials. Peer supporters need training and guidance, emotional support, and well-defined referral mechanisms for clinical questions. Financial support to cover transport reimbursement, modest incentives, and flexible scheduling are essential for long-term viability. Telesupport group expansion will require some technical assistance and family involvement for participants with low literacy levels and well-trained, dedicated moderators. Culturally appropriate materials to engage target audiences effectively may need regular review and updating to ensure they continue to be appropriate. Embedding these elements into programme design would increase both reach and resilience of peer-support interventions within LR care.

Key strengths of this study include its participatory design, contextual diversity, and mixed-methods approach, which captured emotional depth alongside measurable change. Active involvement of individuals affected by LR ensures that each intervention was grounded in lived experience and responsive to local needs. The co-creation processes enhanced relevance, ownership, and trust among participants. The inclusion of both hospital and community clinic settings enabled a nuanced understanding of how peer-support interventions adapt across systems as a perspective rarely addressed in leprosy research.

Nonetheless, limitations must be recognized. First, the small sample size reflects total sampling of all eligible individuals with LR presenting to study sites during the data collection period. Therefore, the small sample size and absence of a control group restrict generalizability and preclude causal inference. The study should be interpreted as an implementation and acceptability pilot rather than an effectiveness trial. During the data collection period, we could not find more LR patients who were willing to participate in the video screening. Some quantitative changes may reflect baseline differences between sites rather than intervention effects. Second, the PV intervention enrolled fewer participants in rural areas due to limited LR cases, which may affect generalizability. Third, family members were interviewed immediately after viewing the video. Therefore, responses likely captured authentic first impressions rather than deeper reflection. As such, reported shifts in attitudes toward providing support should be interpreted as early responses rather than confirmed changes over time. Fourth, the PV was shown once to the participants. The impact of participatory video could be stronger when used in combination with peer discussion rather than as a stand-alone tool[35,41,50–54]. Finally, short follow-up periods captured early changes but may not reflect long-term behavioral outcomes. These caveats highlight the need for larger trials with extended follow-up to assess scalability and durability of impact.

## Conclusions

Our pilot results indicate that all three interventions are promising and result in positive changes in individuals with LR in Indonesia. Each intervention addresses distinct but complementary needs: peer counselling for emotional depth, telesupport group for rapid information exchange, and participatory video for fostering hope for recovery. Outcomes varied by context but consistently indicated improved knowledge, attitude, and practices. The variation across settings illustrates adaptability underscoring that peer-support interventions must align with local infrastructure and culture. Selective and context-driven scale-up, supported by training, digital inclusion, and sustained engagement, offers a realistic pathway to make LR care more person-centered, stigma-sensitive, resilient and thus holistic.

## Acknowledgements

We thank Dr. Elena V. Syurina, Dr. Kevin de Sabbata, and Dr. Brittney S. Mengistu for their contribution to the conceptualization and planning of this pilot. We would like to express our heartfelt gratitude to all the co-implementers and participants who made this study and interventions possible. Your willingness to share experiences, time, and insights has been invaluable, and without your support, we would not have been able to explore new ways to improve the management of leprosy reactions. We are thankful to the leprosy experts across Indonesia, as well as the East Java Provincial Health Office and Bangkalan Health Office, for sharing their knowledge and providing thoughtful input throughout the process. Special thanks go to our dedicated research assistants—Kurnia Siti Mahaniyah, Yusfi Nur Laili Hidayati, and Ummi Kulsum—whose hard work made data collection smooth and successful. We are also grateful to Asiyah Tsabita Maulana for her contribution in developing and analyzing the KAP questionnaire, which enriched this study.

## List of Abbreviations

COVID-19: Coronavirus Disease 2019

ENL: Erythema Nodosum Leprosum

FM: Accompanying Family Member

KAP: Knowledge, attitude, and practices

LR: Leprosy Reactions

MDT: Multidrug therapy

NTD: Neglected tropical diseases

PV: Participatory Video

SARI scale: Stigma Assessment and Reduction of Impact Scale

T1R: Type-1 reactions

WHO: World Health Organization WEMWBS: Warwick Mental Well-Being Scale

## Declarations

### Consent for publication

All participants provided written informed consent for publication and permission to use photographs, except for illiterate participants. For these individuals, the information was conveyed orally and the consent was recorded. Participants involved in interviews, focus group discussions, and video screening consented to the use of anonymised quotes for research and publication purposes. Identifiable information has been removed to ensure confidentiality.

### Availability of data and materials

The datasets generated and analyzed during the current study are not publicly available due to confidentiality involving potentially identifiable information. De-identified excerpts of qualitative data and summary-level quantitative results are available from the corresponding author upon reasonable request for academic purposes. Access requests to the anonymized data (questionnaires, interview guides, and codebook) can be sent to https://doi.org/10.5281/zenodo.20589328. Data will be shared following a formal data-sharing agreement and in accordance with institutional and ethical regulations.

